# Complementary and alternative medicine utilization for malaria prevention: A multi-site community-based cross-sectional study in Ghana

**DOI:** 10.64898/2026.03.29.26349410

**Authors:** Irene A. Kretchy, Augustina Koduah, James-Paul Kretchy, Deborah Atobrah, Hope K. Klobodu, Joseph A. Junior, Nancy Kleponi, Philip T. Mensah, Abidatu Mahama, Michael Opoku-Mireku, Yakubu Alhassan, Afia F.A. Marfo, Mercy Opare-Addo, Harriet A. Bonful, Kwabena F.M. Opuni

## Abstract

**Background:** In many countries, particularly in sub-Saharan Africa, medical pluralism and utilization of multiple therapeutic approaches for managing diseases, including malaria, are common. Ghana’s antimalarial medicine policy has recommended herbal medications for treating uncomplicated malaria. While this is in line with complementary and alternative medicine (CAM) use for the treatment of malaria, exploring CAM for malaria prevention could be an important consideration for public health initiatives towards malaria elimination efforts. This study assessed the prevalence, perceptions, and attitudes on CAM use for malaria prevention in the general population and associated factors.

**Methods:** A community-based cross-sectional analytic survey was conducted among 3064 adult residents sampled between September and November 2023. A multi-stage sampling method was used to select participants from 18 sub-districts drawn from 6 districts and 6 regions in Ghana. Data on CAM use for malaria prevention, attitudes about CAM, perceptions about malaria, and sociodemographic factors were collected. The primary outcome was reported CAM use for malaria prevention within the 12months preceding the survey, measured as a binary outcome. Multiple logistic regression analyses were performed to identify the predictors of CAM use for malaria prevention.

**Results:** A total of 3,064 household respondents were involved in the analysis, with 51.2% (n=1,570) females and a median age of 31 years (IQR: 24-42 years). The use of CAM for malaria prevention in the last 12 months was 31.6% (95% CI: 30.0-33.3%). The most common types of CAM used included botanical/herbal medicine (21.8%), vitamin supplements (12.3%), mineral supplements (10.7%), and spiritual healing/prayers (9.6%). Increased CAM use for malaria prevention was associated with education and perceptions, such as concerns and consequences about malaria. Factors associated with decreased odds of CAM use included formal employment and having a skeptical and indifferent attitude about CAM.

**Conclusion:** Over a third of the population used CAM for malaria prevention in the last year, highlighting its role in public health. Integrating herbal medicine into prevention strategies could enhance community acceptance and help with efforts toward malaria elimination. However, further research is needed to validate clinical efficacy, establish potential drug-herb interactions, and isolate lead compounds for optimized malaria prevention therapy.

## Background

Malaria is a major public health problem, with an estimated 249 million cases reported in 85 malaria-endemic countries [1]. Compared to 2021, there was a 5 million increase in cases for 2022, with most cases and deaths occurring in Sub-Saharan Africa [1–3]. Pregnant women and children under five years old are among the most vulnerable, and in 2022, malaria case incidence was reported as 58 per 1000 for the at-risk population [1].

An estimated 233 million cases of malaria, accounting for 94% of cases globally, were recorded in the WHO African region [1]. To accelerate progress towards malaria elimination and prevention, the WHO Global Strategy for Malaria 2016-2030 envisions a malaria-free world with set targets of a 90% reduction in the global malaria burden by 2030 [4]. Preventive efforts, early diagnosis, and easy access to healthcare and treatment have been prioritized to reduce the incidence of infection, death, and transmission of the parasite [4].

Efforts aimed at malaria prevention have focused on vector control and prevention of malaria in high-risk populations, including pregnant women and children under five years [5]. The use of chemoprophylaxis for travellers entering endemic areas, sleeping under mosquito nets, and using mosquito repellents to prevent a malaria infection, is also utilized [1]. Intermittent preventive therapy in pregnant women (IPTp) and in infancy (IPTi) exemplify chemopreventive measures that benefit maternal, neonatal, and child health, especially in SSA [6, 7]. For some Sahelian communities in SSAs where seasonal transmission is acute, administration of chemoprevention to children between 3 and 59 months is typical [8, 9]. Another malaria preventive initiative is the introduction of the RTS,S, and R21 malaria vaccines for children below five years, whose malaria-related morbidity and mortality rates are high [10, 11].

However, in many countries in SSA, medical pluralism and utilization of multiple therapeutic approaches for the management of diseases such as malaria are common [12, 13]. Both biomedical and CAM are available to patients, either within the formal or informal healthcare system. This pattern accounts for the high utilization of CAM for general healthcare [14–17], with women being more likely to have ever used CAM than men [18, 19]. While biological-based therapies such as herbal medicine are mainly used [20, 21], other modalities like faith-based healing methods (prayer/spirituality) and mind-body therapies (massage, relaxation, meditation, and yoga) have also been reported [22, 23].

The high use has been attributed to the relatively low cost and flexibility in paying for CAM products and services. Other factors such as gender, increased accessibility, perception of the illness, and the perception of CAM as being natural and, therefore, safe and effective compared to conventional therapies have also accounted for their high demand and utilization [19, 24, 25].

The antimalarial medicine policy of Ghana [26], for example, has recommended herbal medications containing *Cryptolepis sanguinolenta, Morinda lucida, Khaya senegalensis, Cassia occidentalis* and *Azadirachta indica* for the treatment of uncomplicated malaria, either as individual medicinal plants or in combination with preventive efforts. While this is in line with the use of herbal medicine for the treatment of malaria over many years, exploring CAM for malaria prevention could be an essential consideration for public health initiatives towards malaria elimination efforts.

While extensive studies have been conducted on the use of CAM modalities to treat malaria [27, 28], little is known about the use of CAM remedies for malaria prevention in the general population and the extent to which perceptions about malaria and attitudes about CAM influence its utilization. Earlier studies in Uganda and Benin have observed CAM use for malaria prevention [29, 30]. In Benin for example, 43.6% of respondents reported using herbal antimalarials for malaria prevention.

In Ghana, two studies provide evidence to document the use of CAM for malaria prevention in Ghana [31, 32]. Dennis Wilmot and colleagues observed the use of CAM products for malaria prevention in Ghana. Similarly, Appiah and colleagues also qualitatively reported the use of CAM for malaria prevention. Both studies, however, do not provide large-scale epidemiological data on the extent of CAM use for malaria prevention the characteristics of users and the associated factors. To the best of our knowledge, this study is potentially the first large-scale epidemiological study assessing the prevalence of CAM use for malaria prevention, and associated sociodemographic, socio-cultural, behavioural factors, including perceptions, and attitudes toward CAM for malaria prevention in the general population based on sex stratification using a nationally representative sample. The study findings will provide baseline information to guide the integration of CAM for malaria prevention to routine malaria prevention strategies and contribute to the national elimination goals.

## Methods

### Study area

The study was conducted in Ghana, a malaria-endemic country with about 30.8 million people and over 5,000 health facilities across public, private, and mission sectors. Healthcare is delivered at primary, secondary, and tertiary levels, with malaria prevention a national priority. CAM is widely used and formally integrated into the health system. CAM services are available in some hospitals, while herbal products are accessible in pharmacies and herbal shops, often without prescriptions, making CAM a culturally accepted approach to malaria prevention.

### Study design

A multi-site community-based analytic cross-sectional study was conducted between September and November 2023 to assess the prevalence, patterns, beliefs, and attitudes towards CAM utilization for malaria prevention and associated factors.

### Study population

The study population included individuals aged 18 years and above who had been living in the community of study for at least three months. Individuals who were sick were excluded from the study.

### Sample size determination

The study employed the Cochrane sample size for hypothesis testing 80% power (thus, *Z*_1−*β*_ = *Z*_1−0.2_ = 0.842), using a conservative 50% prevalence estimate of CAM use for malaria prevention (p=0.50), a margin of error of 5% (thus, e=0.05), at a 0.05 significance level (thus, 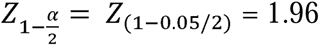). The sample calculation also adjusted for design effect (Deff) using a 0.5 intraclass correlation coefficient (thus, δ=0.5) and 6 clusters (regions) (k=6), and anticipated incomplete record and non-response rate of 10% (thus, r=0.01) based on the formula:

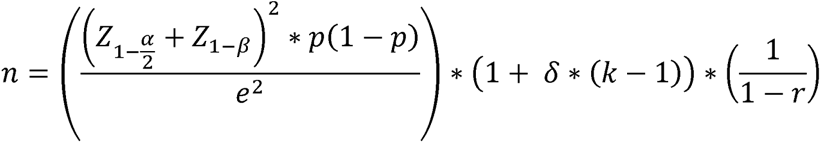

The estimated sample size was 3,057.

### Sampling

A multi-stage sampling approach was used to select the regions, districts, and subdistricts. Households from the subdistricts were then randomly assigned to participate in the study. The study classified the country into three regional zones, *i.e.,* northern, middle, and southern. Two regions were then selected from each zone (Figure 1). According to the 2022 Ghana household and population census report, the region with the largest household population was chosen, and the remaining five regions were selected randomly from the list [33]. One district was randomly chosen from each of the six selected regions. Three sub-districts were randomly selected from each district to sample households (Figure 2).

**Figure 1.**
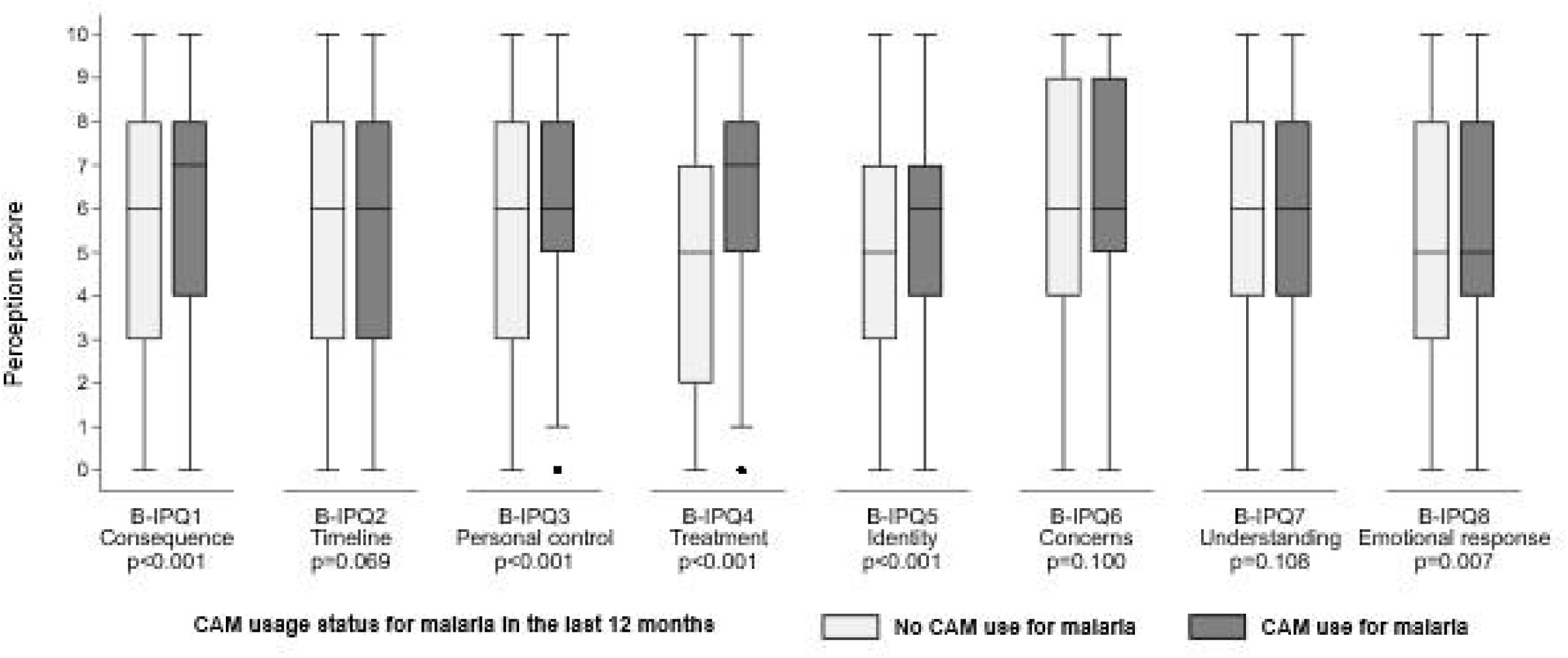
Map of the selected study regions in Ghana

**Figure 2.**
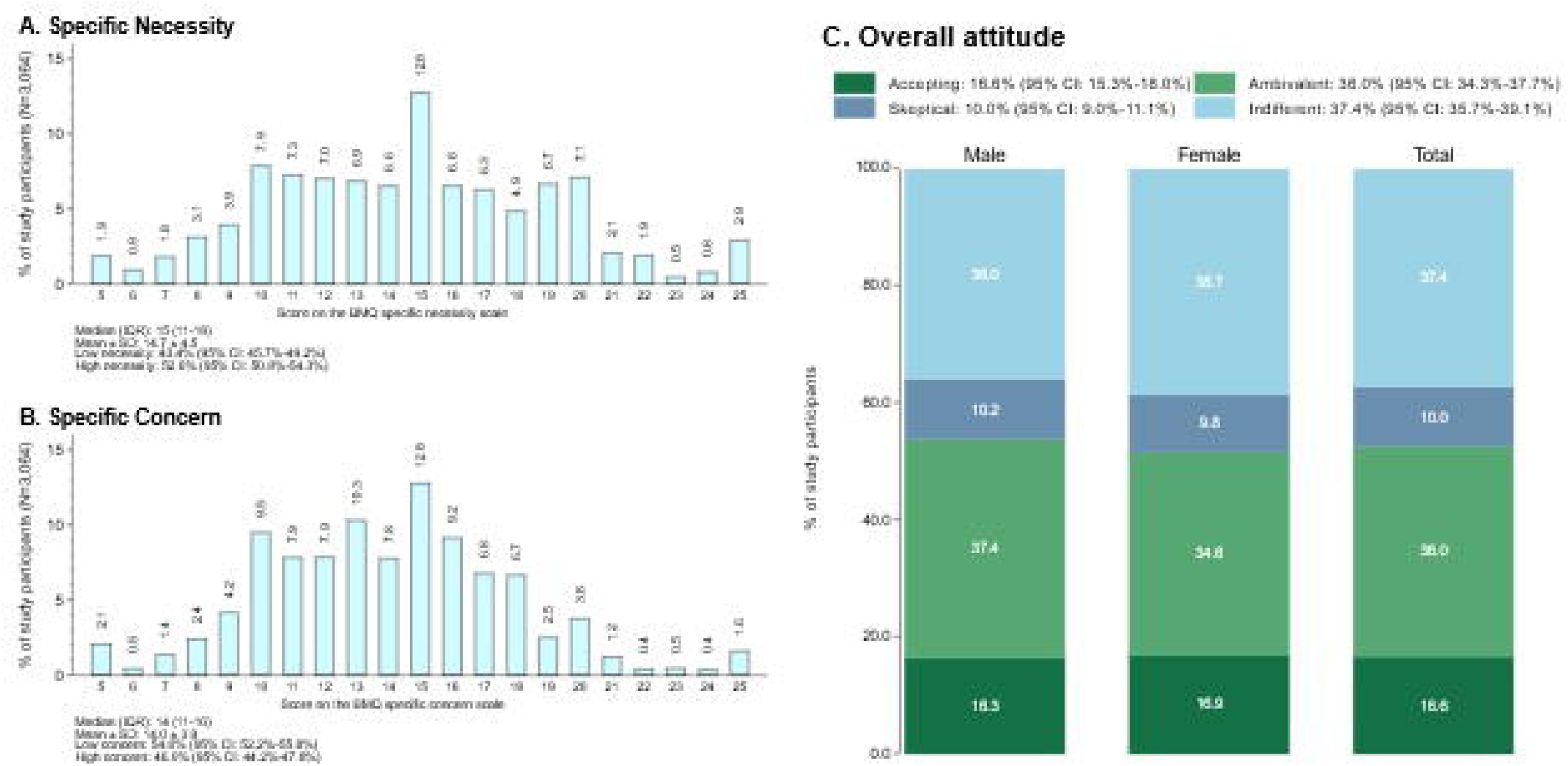
Selected districts and sub-districts for the study

### Sample size allocation

The sample size was proportionally allocated based on the household population of the selected regions obtained from the 2022 Ghana population and housing census [26]. The allocations were as follows: 268 and 162 to the Northern and Upper East regions, respectively, in the northern zones, 930 and 147 to the Ashanti and Western North Region, respectively, in the Middle zone, and 512 and 1,039 to the Central and Greater Accra regions, respectively, in the southern zone (Figure 2). However, the study finally sampled 3,064, achieving a 100.2% rate.

### Data collection and instruments

Information was gathered through a researcher-administered quantitative survey. The data collection tool captured participant details on demographic characteristics, patterns of CAM use for malaria prevention, attitudes towards CAM, and perceptions about malaria. Demographic information such as age, sex, residence, religious affiliation, educational level, employment status, marital status, and household structure was collected. Participants were also asked about their CAM use pattern, including type, dosage, duration, frequency, and reason for use.

Prevalence of CAM use: Respondents were asked about their ever-use of CAM to prevent malaria using a “Yes” or “No” response. A “Yes” response was followed with a question on the last time that CAM was used to prevent malaria. The possible responses to the follow-up question were “Less than a week ago”, “About a month ago”, “About 2-3 months ago”, “4-6 months ago”, “6-12 months ago”, “More than 12 months ago”. The prevalence of CAM use for malaria prevention in the last 12 months was then estimated as the number of responses representing the past 12 months. The questions were adopted from previous studies on CAM use in Ghana [16, 34].

The Beliefs about Medicine Questionnaire (BMQ)-specific was adopted to assess the attitudes toward CAM [27]. Attitudes towards CAM were measured with two sub-scales: necessity for CAM and concern about the potential consequences of CAM for malaria prevention. Each item was scored on a 5-point Likert scale ranging from 1 (strongly disagree) to 5 (strongly agree). The scores of each scale were summed to give a total score. Higher specific-necessity scores indicated stronger attitudes towards the personal need for CAM for malaria prevention. In comparison, higher specific-concerns scores represented stronger concerns about the potential negative effects of CAM. In addition, four attitudinal groups were derived from the scores: Accepting (high necessity, low concerns), Ambivalent (high necessity, high concerns), Indifferent (low necessity, low concerns), and Skeptical (low necessity, high concerns).

The Brief Illness Perception Questionnaire (B-IPQ) was used to assess illness perceptions [35] and was modified to focus on the cognitive and affective illness perceptions/representations towards malaria. The first eight items are scored on an 11-point scale, ranging from 0 to 10. The overall score for the B-IPQ was not computed because each subscale is measured by only one item. In line with previous studies, item 9, an open-ended question on beliefs about causes of illness, was excluded from this study [36, 37].

### Statistical analysis

Field data collected on a hard paper questionnaire was entered into a well-structured Microsoft Excel data entry template with data validation checks and restrictions. Entered data was then imported into Stata MP version 18 (Stata Corp, College Station, TX, USA) for final analysis. Descriptive analysis across all six study regions used frequency and percentages to summarize categorical variables and median and interquartile range to summarize continuous variables. All continuous variables were assessed and confirmed as non-normally distributed using the skewness and kurtosis test. Variation of socio-demographic variable characteristics across the study regions was performed using the Pearson chi-square test for category variables and the Kruskal-Wallis test for continuous variables.

Prevalence of CAM use for malaria prevention and treatment was estimated with the corresponding 95% confidence interval (CI). The pattern of CAM use among respondents who use CAM for malaria prevention was also described across the study regions. The specific CAM types used among respondents for malaria prevention and treatment were also represented on a bar chart.

Individual items on the BMQ were described across the study participants. The four attitudinal groups obtained from the BMQ scale were described, and their association with CAM use for malaria was assessed using the Pearson chi-square test, which was presented on a bar chart.

Each of the 8 items on the B-IPQ was summarized using the median and interquartile range. In line with previous studies, the overall score for the B-IPQ was not computed [16, 29]. The distribution of the scores from each individual item on the B-IPQ scale was compared between users and non-users of CAM for malaria prevention and displayed using a boxplot. The equality of medians was assessed using the Wilcoxon rank sum test.

The binary logistic regression model was then used to estimate the crude and adjusted odds of CAM use for malaria prevention and treatment among the study participants. For the multiple-adjusted binary logistic regression model, except for the excluded B-IPQ items, all other variables were included in the model due to high multicollinearity between the items of the B-IPQ scale. Next, separate models were fitted for each item on the B-IPQ scale, adjusting for the effect of all other socio-demographic characteristics, including the attitudinal group. The corresponding 95% CI and p-value for all odds ratios were estimated. Multicollinearity was assessed using the variance inflation factor, which obtained a mean VIF of 2.64 (range: 1.13-5.77), within the recommended range of less than 10. All statistical analyses were considered significant at 0.05 alpha level.

## Results

Table 1 describes the sociodemographic characteristics of the participants by sex. A total of 3064 participants were involved in the study, with 51.2% (n=1,570) females. The median age was 31 years (24-42), with 7.6% (n=234) older than 60 years. Overall, the majority (76.5%, n= 2345) of the participants were Christians, with at least 7 out of every 10 participants in 5 regions being Christians, excluding the Upper East region, where 75.3% were Muslims. Almost half (48.5%, n=1487) of the participants were single. A minority (7.8%, n=240) of the participants had not benefited from any education. Among the participants, 68.8% were employed in the formal or informal sectors of the economy. One in every five participants (20.2%, n=619) had an existing health condition. The most common health conditions reported were hypertension (11.4%), peptic ulcer (7.5%), diabetes (6.1%), asthma (5.8%) and colds (5.1%). Significant variations between male and female respondents were observed in some of the socio-demographic characteristics, including region (p<0.001), age group (p=0.006), religion (p<0.001), marital status (p=0.006), highest education (p<0.001), employment status (p<0.001), ethnicity (p<0.001), household size (p<0.001), number of children (p<0.001), and history of existing medical condition (p=0.040), in survey (p<0.05) (Table 1).

**Table 1.**
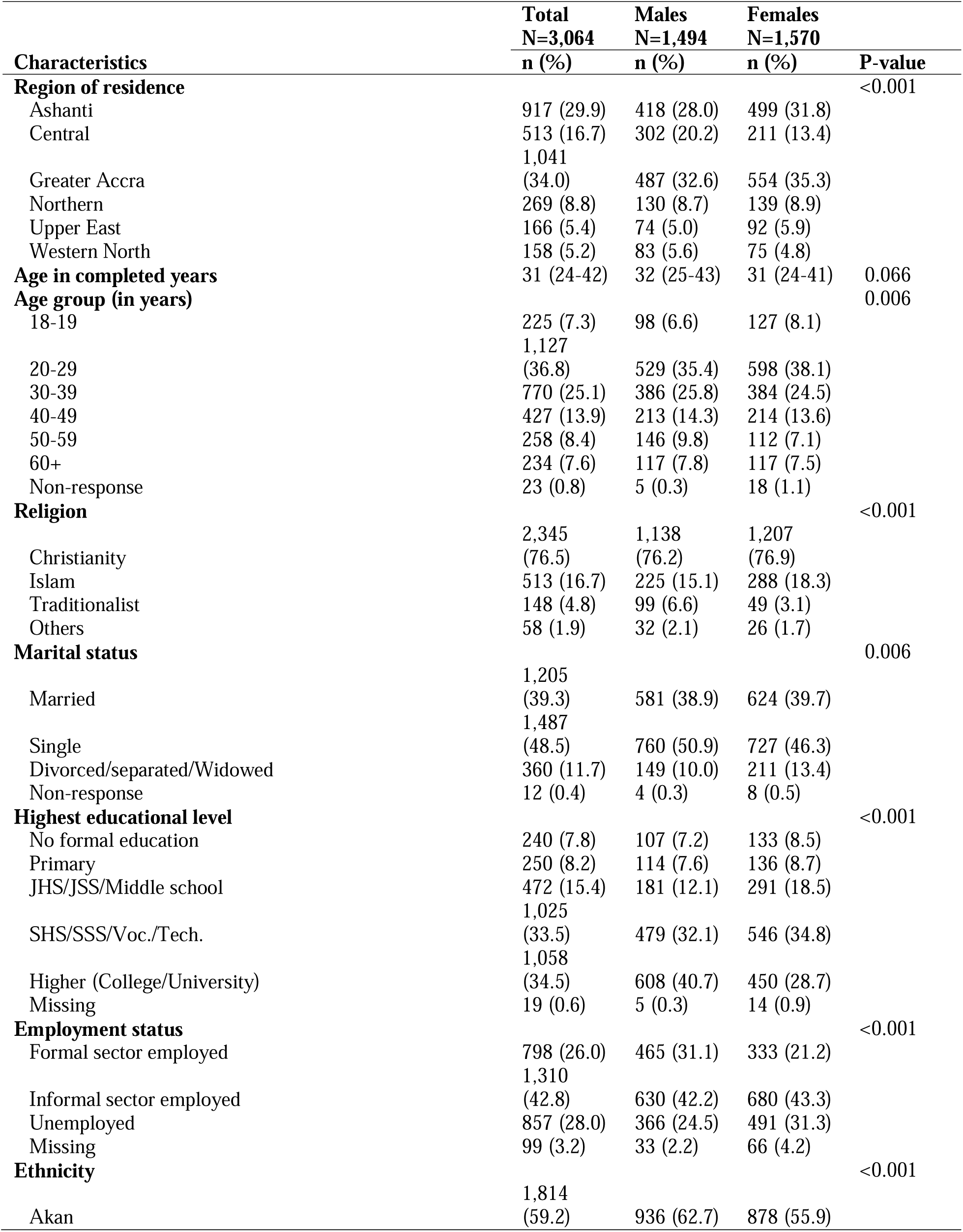
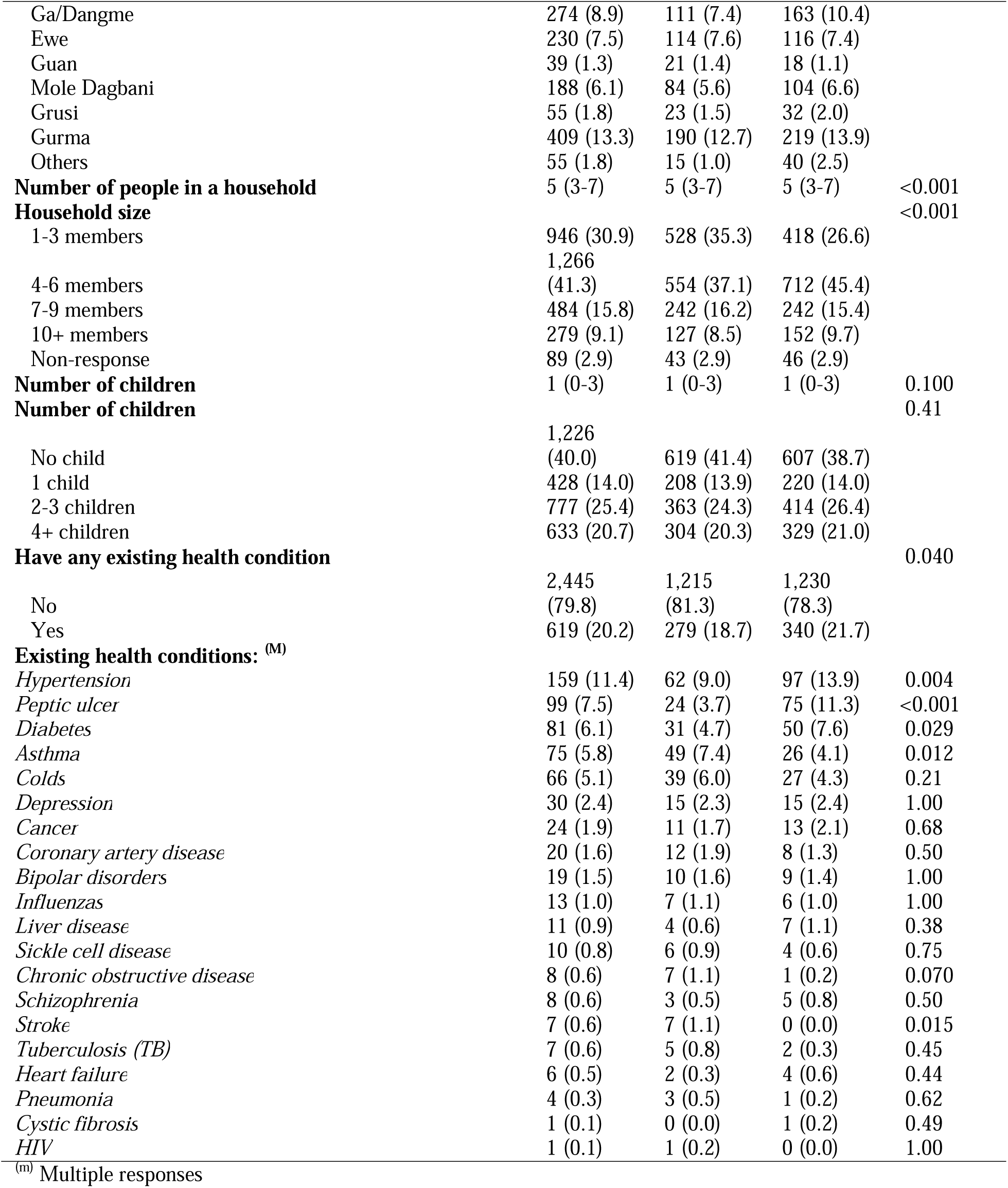
Demographic characteristics of study respondents by sex.

### Prevalence and pattern of use of CAM in the last 12 months for malaria prevention among study participants in Ghana

Overall, the proportion of participants who had ever used CAM to prevent malaria infection was 31.6% (95% CI: 30.0-33.3%). CAM use for malaria prevention in the last 12 months was 32.9% (95% CI: 30.5-35.3%) among males and 30.4% (95% CI: 28.2-32.8%) among females (p=0.150). CAM use for malaria in the last 12 months was highest in the Central (55.6%) and Northern (53.9%) regions and lowest in the Greater Accra (15.6%) region. Within regions, CAM use for malaria prevention in the last 12 months did not significantly vary between males and females (p>0.05) (Figure 3).

**Figure 3.**
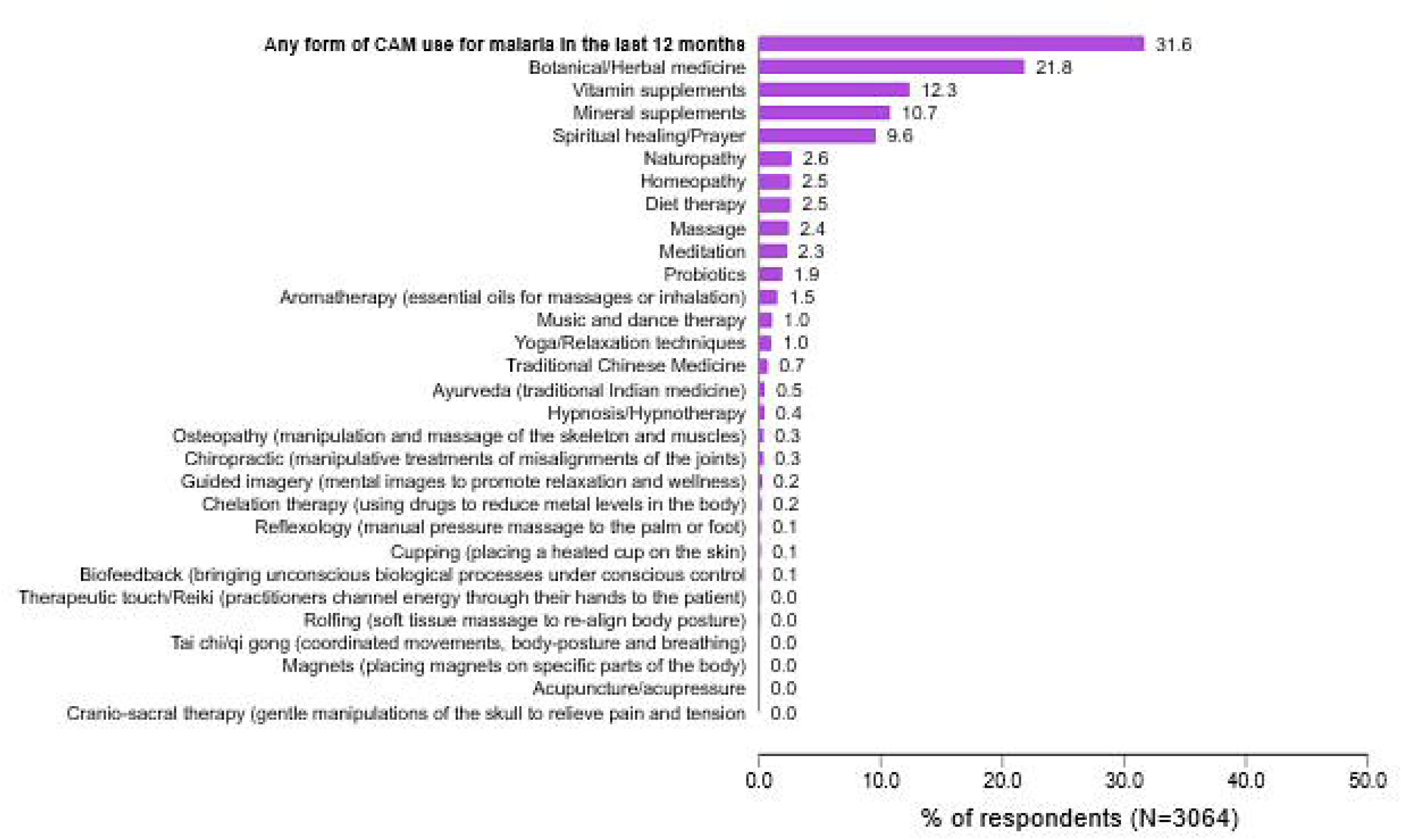
Prevalence of CAM use for malaria prevention and treatment

In decreasing order, the most common forms of CAM for malaria prevention among all participants were herbal medicines (21.8%), vitamin supplements (12.3%), mineral supplements (10.7%), and spiritual healing/prayers (9.6%) (Figure 4).

**Figure 4.**
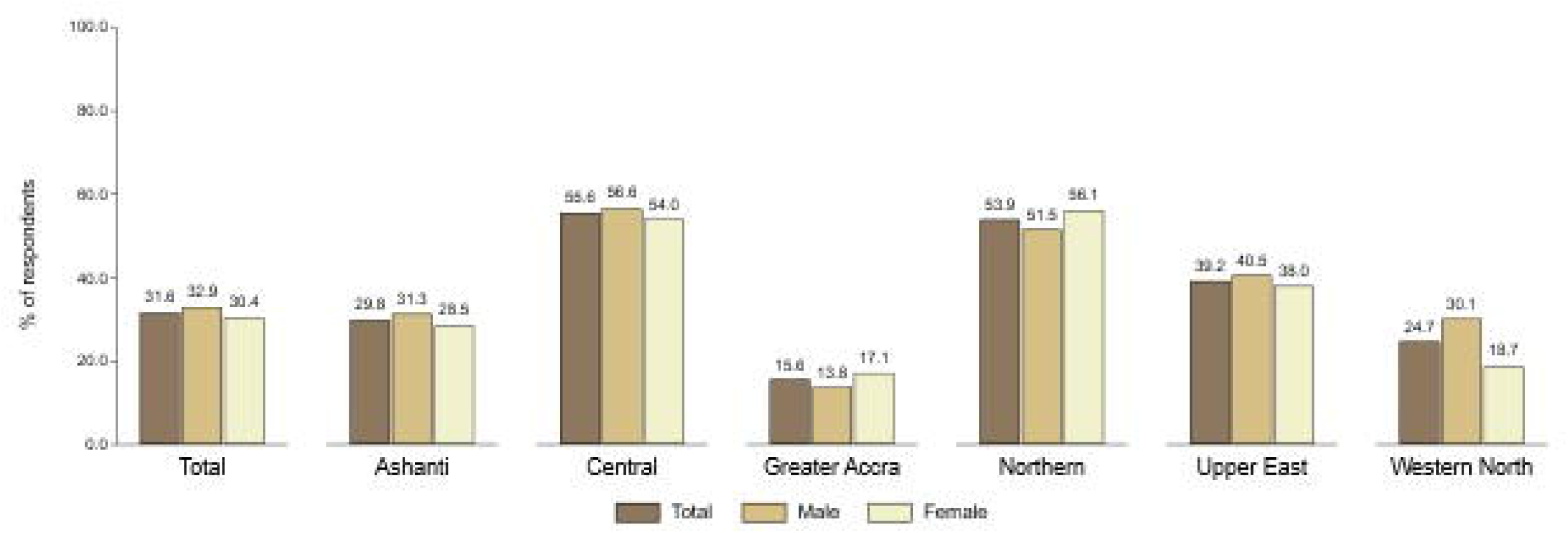
Specific CAM usage among CAM users for malaria prevention

A summary of the pattern of use of CAM to reduce malaria risk among those who use CAM for malaria prevention is summarised in Table 2. While 17.9% of the participants had relied on CAM to prevent malaria infection for over 10 years, a majority (60.5%, n=587) reportedly used CAM within the past 12 months. Most participants (81.7%, n=793) used CAM to reduce their malaria risk within six months before the survey. Most (81.7%, n=791) of the participants took the CAM for malaria prevention either once, twice, or thrice a day. Participants relied mainly on the practitioner’s advice (52.4%, n=508), product label instructions (41.3%, n=400), and personal choice (32.8%, n=318) for direction on how to take the CAM to prevent malaria. Most participants (48.9% (n=474) relied on products with Ghana FDA registration. Except for 48.8% (n=473) of participants who did not mix their CAM for malaria prevention with other medications, the rest reportedly added orthodox drugs to the CAM for malaria prevention. Headaches (30.0%), fatigue (23.6%), and dizziness (20.0%) were the top three side effects experienced by the participants after intake of malaria-preventing CAM products. The various pattern of CAM use for malaria prevention significantly varies between males and females (p<0.001) (Table 2).

**Table 2.** Pattern of CAM usage for malaria prevention among respondents who use CAM for malaria prevention by region.

| Characteristics | Total<br>N=969<br>n (%) | Male<br>N=491 | Female<br>N=478 | P-value |
| --- | --- | --- | --- | --- |
| **Duration of CAM used to prevent malaria** |  |  |  | 0.005 |
| Less than 6 months | 295 (30.4) | 170 (34.6) | 125 (26.2) |  |
| 6-12 months | 292 (30.1) | 134 (27.3) | 158 (33.1) |  |
| 1-5 years | 147 (15.2) | 64 (13.0) | 83 (17.4) |  |
| 6-9 years | 58 (6.0) | 28 (5.7) | 30 (6.3) |  |
| 10 or more years | 173 (17.9) | 95 (19.3) | 78 (16.3) |  |
| Missing | 4 (0.4) | 0 (0.0) | 4 (0.8) |  |
| **Last time CAM was used to prevent malaria** |  |  |  | 0.046 |
| Less than a week ago | 145 (15.0) | 75 (15.3) | 70 (14.6) |  |
| About a month ago | 281 (29.0) | 131 (26.7) | 150 (31.4) |  |
| About 2-3 months ago | 190 (19.6) | 85 (17.3) | 105 (22.0) |  |
| 4-6 months ago, | 177 (18.3) | 101 (20.6) | 76 (15.9) |  |
| 6-12 months ago, | 176 (18.2) | 99 (20.2) | 77 (16.1) |  |
| **Frequency of CAM use to prevent malaria** |  |  |  | 0.001 |
| Three or more times daily | 239 (24.7) | 140 (28.5) | 99 (20.7) |  |
| Two times daily | 283 (29.2) | 127 (25.9) | 156 (32.6) |  |
| Once a Day | 269 (27.8) | 145 (29.5) | 124 (25.9) |  |
| 2-3 times a week | 55 (5.7) | 33 (6.7) | 22 (4.6) |  |
| 4-6 times a week | 52 (5.4) | 16 (3.3) | 36 (7.5) |  |
| Twice a month | 19 (2.0) | 10 (2.0) | 9 (1.9) |  |
| Once a month | 12 (1.2) | 5 (1.0) | 7 (1.5) |  |
| Once every 3 months | 5 (0.5) | 2 (0.4) | 3 (0.6) |  |
| Occasionally | 30 (3.1) | 13 (2.6) | 17 (3.6) |  |
| Missing | 5 (0.5) | 0 (0.0) | 5 (1.0) |  |
| **How did you decide on the frequency of therapy, during your last use of CAM for malaria prevention** |  |  |  |  |
| Followed product label instructions | 400 (41.3) | 232 (47.3) | 168 (35.1) | <0.001 |
| Followed practitioner's instructions | 508 (52.4) | 283 (57.6) | 225 (47.1) | <0.001 |
| Decided myself | 318 (32.8) | 151 (30.8) | 167 (34.9) | 0.17 |
| Media (e.g. TV, Radio, Newspapers) | 54 (5.6) | 24 (4.9) | 30 (6.3) | 0.35 |
| Friends/Relatives | 205 (21.2) | 91 (18.5) | 114 (23.8) | 0.043 |
| Social media (e.g. WhatsApp, Facebook, Twitter etc.) | 17 (1.8) | 11 (2.2) | 6 (1.3) | 0.24 |
| Internet | 16 (1.7) | 7 (1.4) | 9 (1.9) | 0.58 |
| **Source of CAM for malaria prevention** |  |  |  |  |
| Pharmacy | 312 (32.2) | 188 (38.3) | 124 (25.9) | <0.001 |
| Chemical store | 27 (2.8) | 15 (3.1) | 12 (2.5) | 0.61 |
| Herbal medicine store | 177 (18.3) | 90 (18.3) | 87 (18.2) | 0.96 |
| Practitioner | 80 (8.3) | 32 (6.5) | 48 (10.0) | 0.046 |
| Friends/Family | 161 (16.6) | 78 (15.9) | 83 (17.4) | 0.54 |
| Internet/online | 7 (0.7) | 1 (0.2) | 6 (1.3) | 0.053 |
| Raw materials/home-made | 344 (52.7) | 166 (48.4) | 178 (57.4) | 0.021 |
| Finished products | 578 (70.8) | 303 (71.0) | 275 (70.7) | 0.93 |
| **Products registered by the FDA** |  |  |  | 0.47 |
| I always use registered product | 474 (48.9) | 253 (51.5) | 221 (46.2) |  |
| I sometimes use unregistered product | 88 (9.1) | 40 (8.1) | 48 (10.0) |  |
| I never use registered product | 71 (7.3) | 37 (7.5) | 34 (7.1) |  |
| I am not sure; I just use them | 322 (33.2) | 155 (31.6) | 167 (34.9) |  |
| Missing | 14 (1.4) | 6 (1.2) | 8 (1.7) |  |
| **Informs medical doctor/pharmacist about use of CAM for malaria prevention or treatment** |  |  |  | 0.052 |
| Yes | 437 (45.1) | 216 (44.0) | 221 (46.2) |  |
| No | 498 (51.4) | 264 (53.8) | 234 (49.0) |  |
| Missing | 34 (3.5) | 11 (2.2) | 23 (4.8) |  |
| **Normally mix orthodox medication with CAM for malaria prevention or treatment** |  |  |  | 0.015 |
| Yes, for both prevention and treatment | 262 (27.0) | 133 (27.1) | 129 (27.0) |  |
| Yes, for prevention only | 92 (9.5) | 39 (7.9) | 53 (11.1) |  |
| Yes, for treatment only | 126 (13.0) | 50 (10.2) | 76 (15.9) |  |
| No | 473 (48.8) | 260 (53.0) | 213 (44.6) |  |
| Missing | 16 (1.7) | 9 (1.8) | 7 (1.5) |  |
| **Perceived effectiveness of CAM for malaria prevention** |  |  |  | 0.054 |
| Extremely effective | 241 (24.9) | 108 (22.0) | 133 (27.8) |  |
| Very effective | 514 (53.0) | 284 (57.8) | 230 (48.1) |  |
| Slightly effective | 147 (15.2) | 69 (14.1) | 78 (16.3) |  |
| Not effective | 54 (5.6) | 25 (5.1) | 29 (6.1) |  |
| Problematic | 3 (0.3) | 2 (0.4) | 1 (0.2) |  |
| Missing | 10 (1.0) | 3 (0.6) | 7 (1.5) |  |
| **Side effects experienced using CAM for malaria prevention: <sup>(M)</sup>** |  |  |  |  |
| *Headache* | 291 (30.0) | 135 (27.5) | 156 (32.6) | 0.092 |
| *Fatigue* | 229 (23.6) | 107 (21.8) | 122 (25.5) | 0.17 |
| *Dizziness* | 194 (20.0) | 83 (16.9) | 111 (23.2) | 0.016 |
| *Vomiting* | 144 (14.9) | 56 (11.4) | 88 (18.4) | 0.003 |
| *Nausea* | 113 (11.7) | 50 (10.2) | 63 (13.2) | 0.16 |
| *Diarrhoea* | 90 (9.3) | 43 (8.8) | 47 (9.8) | 0.58 |
| *Pain* | 67 (6.9) | 36 (7.3) | 31 (6.5) | 0.62 |
| *Allergic reactions* | 56 (5.8) | 41 (8.4) | 15 (3.1) | <0.001 |
| *Dermatitis* | 20 (2.1) | 8 (1.6) | 12 (2.5) | 0.37 |
| *Skin burns* | 19 (2.0) | 13 (2.6) | 6 (1.3) | 0.16 |
| *Joint injury* | 7 (0.7) | 5 (1.0) | 2 (0.4) | 0.45 |
| *Kidney toxicity* | 6 (0.6) | 1 (0.2) | 5 (1.0) | 0.12 |
| *Bruising* | 4 (0.4) | 3 (0.6) | 1 (0.2) | 0.62 |
| *Spinal injury* | 3 (0.3) | 3 (0.6) | 0 (0.0) | 0.25 |
| *Heart toxicity* | 2 (0.2) | 1 (0.2) | 1 (0.2) | 1.00 |
| *Hepatitis* | 2 (0.2) | 1 (0.2) | 1 (0.2) | 1.00 |
| *Liver toxicity* | 1 (0.1) | 0 (0.0) | 1 (0.2) | 0.49 |

### Attitude and Perception towards CAM for Malaria prevention

#Figure 5 shows that the median score for specific necessity towards CAM for malaria prevention was 15 (IQR: 11-18). More than half (52.6%, 95% CI: 50.8 - 54.3) had a high necessity for malaria prevention. Low concern about CAM for malaria prevention was 54.0% (95% CI: 52.2-55.8). In summary, while 1 out of every 10 (10.0%) participant was skeptical about using CAM to reduce malaria risk, 16.6% accepted CAM to prevent a malaria infection, 36.0% were ambivalent, and 37.4% were indifferent. Similar patterns of attitude toward CAM use to prevent malaria infection were observed independently for both males and females (Figure 5).

**Figure 5.**
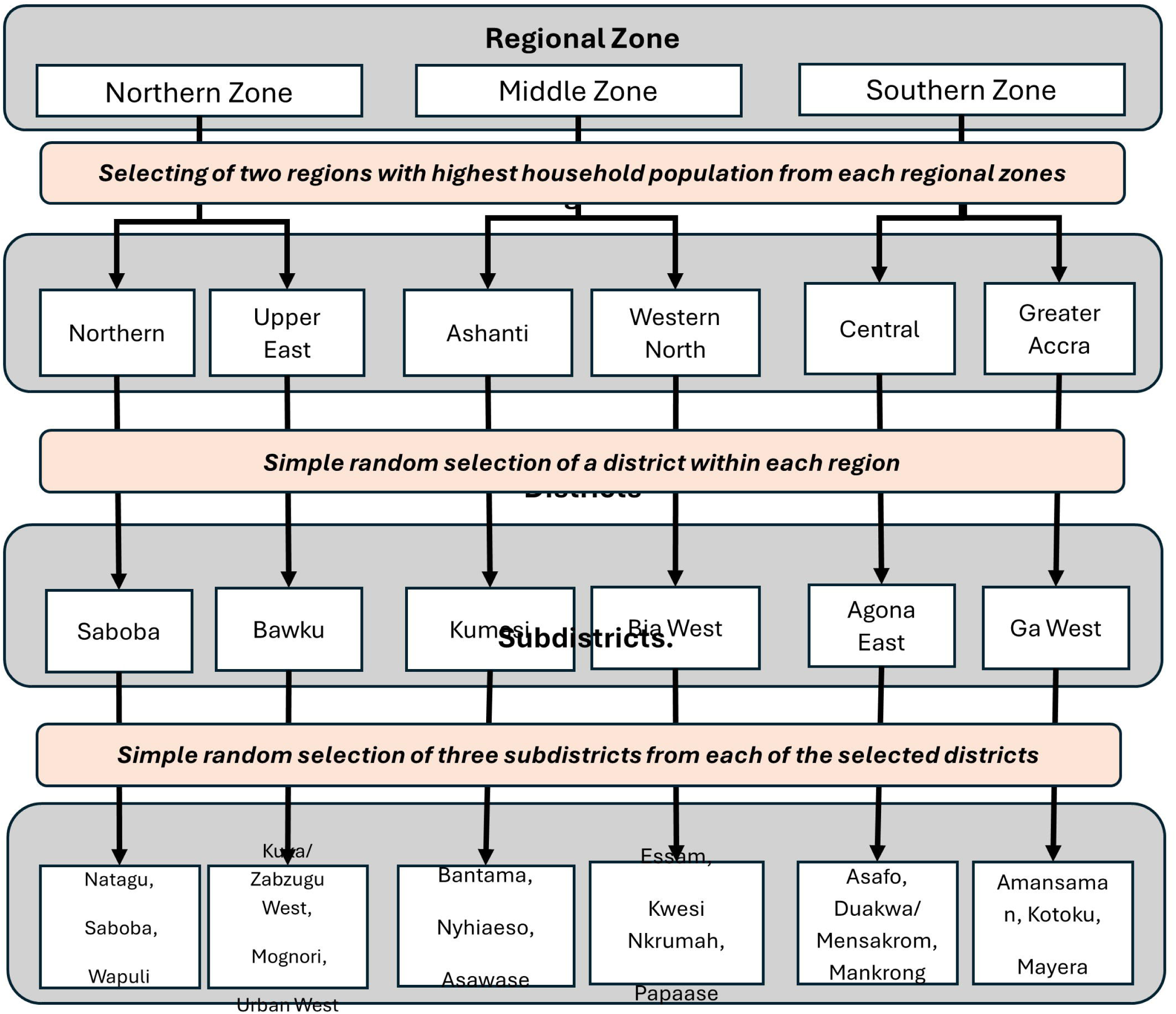
Attitude towards CAM usage for malaria prevention

### Perception of study participants towards malaria by region

Table 3 summarises the participants’ perceptions towards malaria prevention among the respondents by sex. Except for emotional response and treatment, a high median score of 6 was recorded for all other perception variables (Table 3).

**Table 3.** Perception of study participants towards malaria by sex.

| Item<br>code | Perception | Short<br>description | Total<br>N=3,0<br>64 | Males<br>N=1,4<br>94 | Femal<br>es<br>N=1,5<br>70 | P-value |
| --- | --- | --- | --- | --- | --- | --- |
|  |  |  | Media<br>n<br>(IQR) | Media<br>n<br>(IQR) | Media<br>n<br>(IQR) |  |
| **B-IPQ**<br>**1** | How much does malaria affect your life? | Consequence | 6 (3-8) | 6 (3-8) | 6 (3-8) | 0.750 |
| **B-IPQ**<br>**2** | How long do you think malaria will continue? | Timeline | 6 (3-8) | 6 (3-8) | 6 (3-8) | 0.600 |
| **B-IPQ**<br>**3** | How much control do you feel you have over malaria? | Personal control | 6 (4-8) | 6 (4-8) | 6 (3-8) | 0.004 |
| **B-IPQ**<br>**4** | How much do you think CAM can help with malaria? | Treatment | 5 (3-8) | 5 (3-8) | 5 (3-8) | 0.310 |
| **B-IPQ**<br>**5** | How much do you experience symptoms of malaria? | Identity | 6 (3-7) | 5 (3-7) | 6 (3-7) | 0.570 |
| **B-IPQ**<br>**6** | How concerned are you about malaria? | Concerns | 6 (4-9) | 6 (4-9) | 6 (4-9) | 0.380 |
| **B-IPQ**<br>**7** | How well do you feel you understand malaria? | Understanding | 6 (4-8) | 6 (4-8) | 6 (4-8) | 0.032 |
| **B-IPQ**<br>**8** | How much does malaria affect you emotionally? | Emotional response | 5 (3-8) | 5 (3-7) | 5 (3-8) | 0.660 |

### Bivariate association between Demographic characteristics of respondents and CAM usage for malaria prevention and treatment

Bivariate Chi square analysis of the association between sociodemographic factors, and attitudes are presented in Table 4. All the sociodemographic factors, including age, sex, religion, and marital status, as well as the presence of comorbidities, were significantly associated with CAM for malaria prevention. Furthermore, attitude was significantly associated with the use of CAM for the prevention of malaria (P<0.05). A similar significance level was observed when the respondents were stratified by sex (Table 4).

**Table 4.**
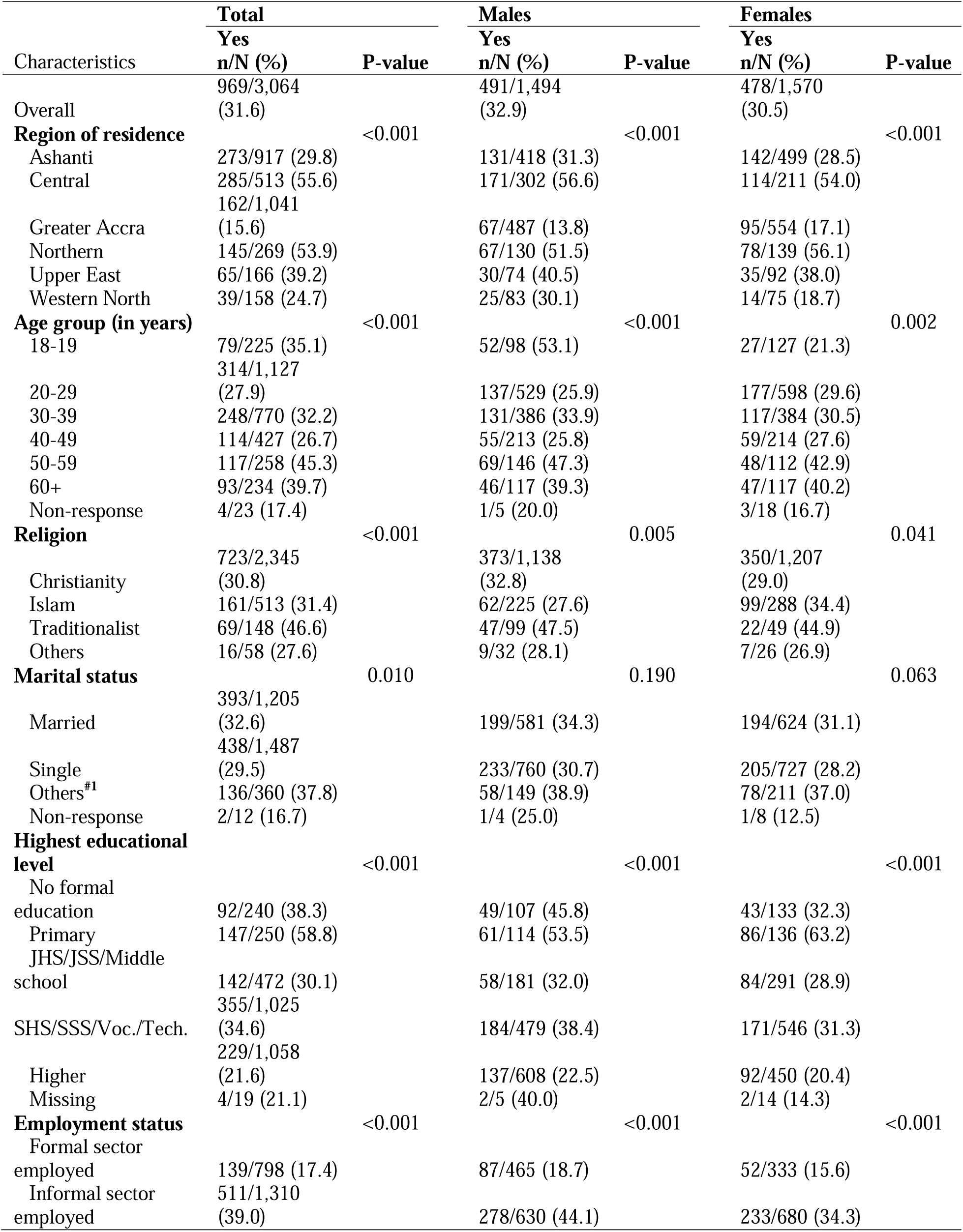
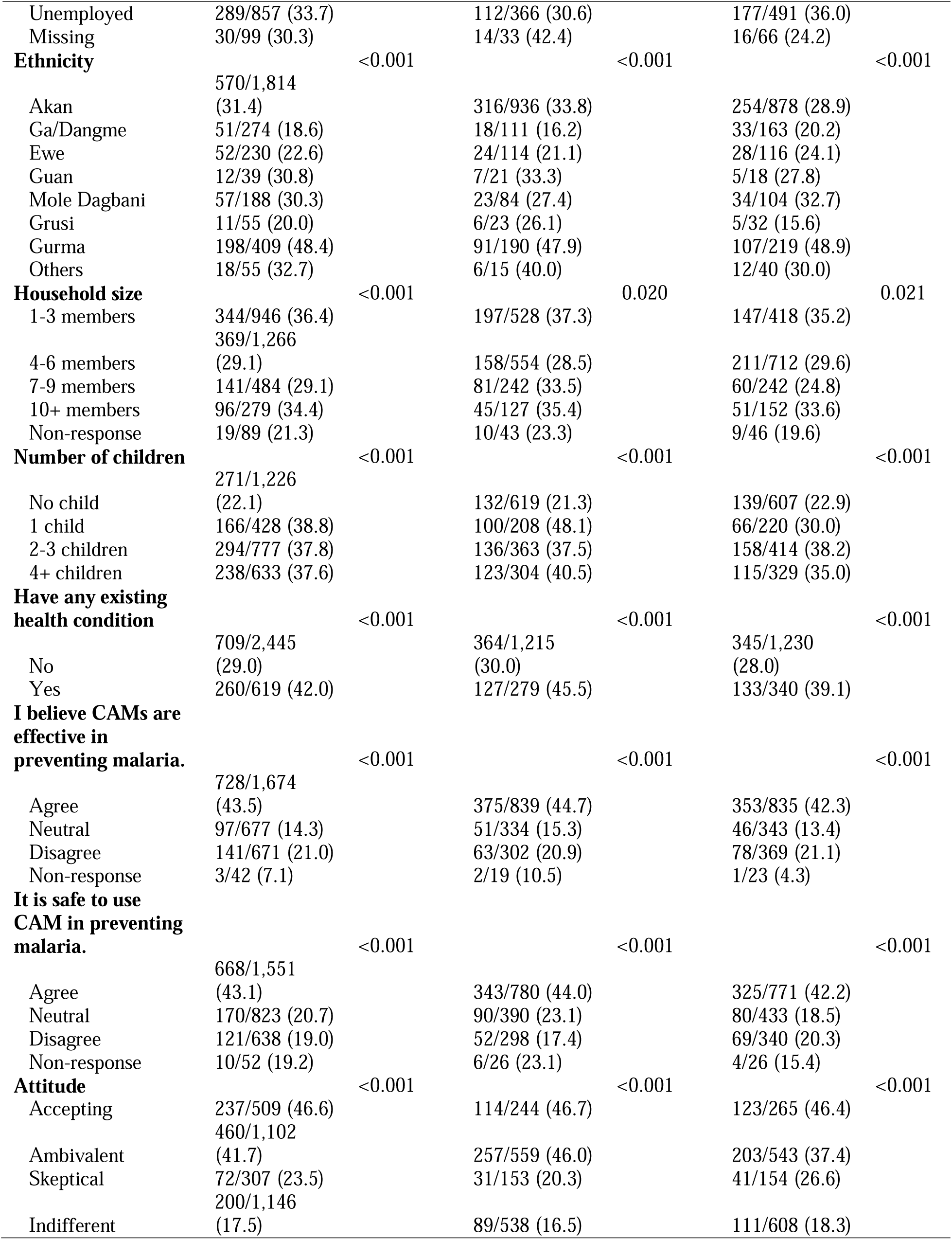
Bivariate association between Demographic characteristics of respondents and CAM usage for malaria prevention and treatment.

CAM use for malaria prevention in the last 12 months was highest among respondents whose attitude towards CAM use for malaria was accepting (46.6%) or ambivalent (41.7%) compared to skeptical (23.5%) and those who were indifferent (17.5%) (Table 4).

The bivariate association between CAM use for malaria in the last 12 months and perception was significant in terms of consequences (p<0.001), personal control (p<0.001), treatment (p<0.001), identity (p<0.001) and emotional response (p=0.007) (Figure 6).

**Figure 6.**
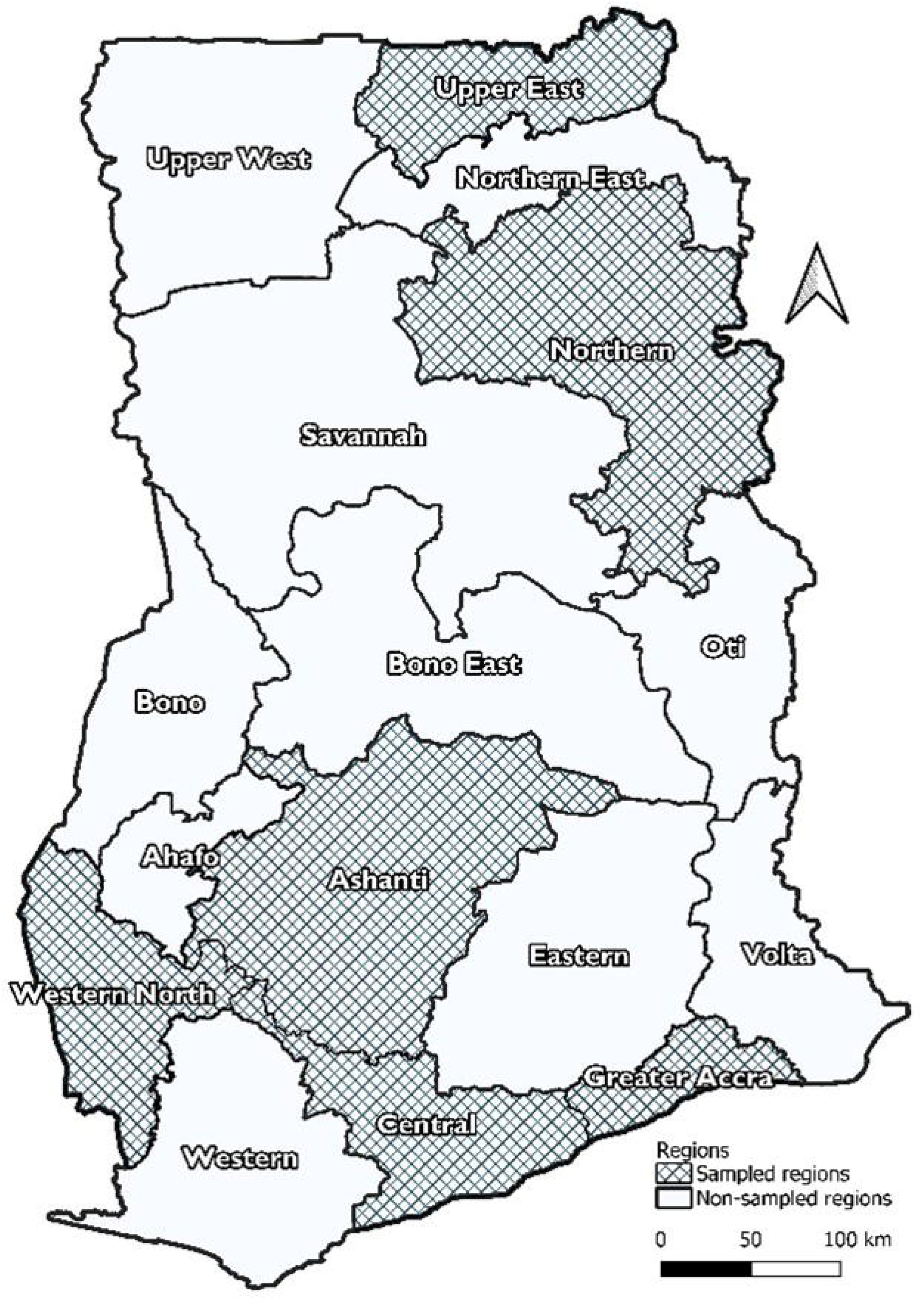
Bivariate association between CAM usage for malaria and perception of malaria

### Binary logistic regression model of factors associated with CAM usage for malaria prevention and treatment

Univariable logistic regression analyses revealed that sex, educational level, employment status, number of children, and presence of comorbidities, as well as perceptions and attitudes, were associated with the use of CAM for malaria risk reduction (p<005). In the adjusted model, however, perceptions and attitudes significantly predicted CAM for malaria prevention (p<0.05).

Relative to participants from the Greater Accra region, the use of CAM for malaria prevention in the last 12 months was about 3 times in the Ashanti region (aOR: 2.96, 95% CI: 2.21-3.98, p<0.001), about 4 times in the Central (aOR: 3.72, 95% CI: 2.50-5.53, p<0.001) and Northern (aOR: 3.88, 95% CI: 2.17-6.93, p<0.001) and over 6 times in the Upper East (aOR: 6.23, 95% CI: 3.47-11.17, p<0.001).

Relative to married respondents, CAM usage for malaria prevention in the last 12 months was 32% less among the divorced/separated/widowed (aOR: 0.68, 95% CI: 0.49-0.94, p=0.021). Relative to those without formal education, the use of CAM for malaria in the last 12 months was higher among those with primary (aOR: 2.33, 95% CI: 1.49-3.63, p<0.001), secondary (aOR: 1.69, 95% CI: 1.12-2.56, p=0.013) and higher (aOR: 2.05, 95% CI: 1.29-3.25, p=0.002).

Relative to those unemployed, CAM used for malaria prevention in the last 12 months was 44% less among those formally employed (aOR: 0.56, 95% CI: 0.40-0.80).

Participants who were skeptical about using CAM to prevent malaria were 44% less likely to use CAM to prevent malaria infection than their comparators who accepted CAM for malaria prevention (aOR: 0.56, 95% CI: 0.37-0.85, p=0.006). Participants who were indifferent towards using CAM to prevent malaria were 54% less likely to use CAM to prevent malaria than those who accepted CAM for malaria prevention (aOR: 0.49, 95% CI: 0.35-0.68, p<0.001).

In terms of perception of malaria, the odds of CAM use for malaria prevention in the last 12 months increase with an increase in perception of CAM effectiveness for malaria treatment (aOR: 1.18, 95% CI: 1.12-1.23) (Table 5).

**Table 5.** Binary logistic regression model of factors associated with CAM usage for malaria prevention and treatment.

| Characteristics | Binary logistic regression model |  |  |  |
| --- | --- | --- | --- | --- |
|  | Unadjusted model |  | Adjusted model |  |
|  | cOR [95% CI] | P-value | aOR [95% CI] | P-value |
| **Sex of participants** |  |  |  | 0.989<sup>#</sup> |
| Males | 1.00 [reference] |  | 1.00 [reference] |  |
| Females | 0.89 [0.77, 1.04] | 0.150 | 1.00 [0.83, 1.21] | 0.989 |
| **Region of residence** |  |  |  | <0.001<sup>#</sup> |
| Ashanti | 2.30 [1.85, 2.86] | <0.001 | 2.96 [2.21, 3.98] | <0.001 |
| Central | 6.78 [5.33, 8.64] | <0.001 | 3.72 [2.50, 5.53] | <0.001 |
| Greater Accra | 1.00 [reference] |  | 1.00 [reference] |  |
| Northern | 6.34 [4.74, 8.50] | <0.001 | 3.88 [2.17, 6.93] | <0.001 |
| Upper East | 3.49 [2.45, 4.97] | <0.001 | 6.23 [3.47, 11.17] | <0.001 |
| Western North | 1.78 [1.19, 2.65] | 0.005 | 1.25 [0.76, 2.06] | 0.372 |
| **Age group (in years)** |  |  |  | 0.003<sup>#</sup> |
| 18-19 | 1.00 [reference] |  | 1.00 [reference] |  |
| 20-29 | 0.71 [0.53, 0.97] | 0.029 | 0.83 [0.58, 1.20] | 0.322 |
| 30-39 | 0.88 [0.64, 1.20] | 0.415 | 0.87 [0.58, 1.31] | 0.497 |
| 40-49 | 0.67 [0.48, 0.95] | 0.026 | 0.85 [0.52, 1.40] | 0.530 |
| 50-59 | 1.53 [1.06, 2.21] | 0.023 | 1.54 [0.91, 2.58] | 0.105 |
| 60+ | 1.22 [0.83, 1.78] | 0.306 | 0.97 [0.55, 1.73] | 0.921 |
| Non-response | 0.39 [0.13, 1.18] | 0.096 | 0.18 [0.05, 0.65] | 0.009 |
| **Religion** |  |  |  | 0.490<sup>#</sup> |
| Christianity | 1.00 [reference] |  | 1.00 [reference] |  |
| Islam | 1.03 [0.84, 1.26] | 0.806 | 0.91 [0.68, 1.21] | 0.503 |
| Traditionalist | 1.96 [1.40, 2.74] | <0.001 | 1.36 [0.85, 2.16] | 0.196 |
| Others | 0.85 [0.48, 1.53] | 0.597 | 0.98 [0.48, 1.97] | 0.946 |
| **Marital status** |  |  |  | 0.042<sup>#</sup> |
| Married | 1.00 [reference] |  | 1.00 [reference] |  |
| Single | 0.86 [0.73, 1.02] | 0.078 | 1.10 [0.82, 1.48] | 0.511 |
| Divorced/separated/Widowed | 1.25 [0.98, 1.60] | 0.070 | 0.68 [0.49, 0.94] | 0.021 |
| Non-response | 0.41 [0.09, 1.90] | 0.255 | 0.55 [0.16, 1.88] | 0.343 |
| **Highest educational level** |  |  |  | <0.001<sup>#</sup> |
| No formal education | 1.00 [reference] |  | 1.00 [reference] |  |
| Primary | 2.30 [1.60, 3.30] | <0.001 | 2.33 [1.49, 3.63] | <0.001 |
| JHS/JSS/Middle school | 0.69 [0.50, 0.96] | 0.027 | 1.11 [0.73, 1.69] | 0.613 |
| SHS/SSS/Voc./Tech. | 0.85 [0.64, 1.14] | 0.281 | 1.69 [1.12, 2.56] | 0.013 |
| Higher (College/University) | 0.44 [0.33, 0.60] | <0.001 | 2.05 [1.29, 3.25] | 0.002 |
| Missing | 0.43 [0.14, 1.33] | 0.143 | 1.38 [0.47, 4.06] | 0.564 |
| **Employment status** |  |  |  | <0.001<sup>#</sup> |
| Formal sector employed | 0.41 [0.33, 0.52] | <0.001 | 0.56 [0.40, 0.80] | 0.001 |
| Informal sector employed | 1.26 [1.05, 1.50] | 0.013 | 1.27 [0.96, 1.69] | 0.089 |
| Unemployed | 1.00 [reference] |  | 1.00 [reference] |  |
| Missing | 0.85 [0.54, 1.34] | 0.495 | 0.82 [0.48, 1.40] | 0.469 |
| **Ethnicity** |  |  |  | 0.008<sup>#</sup> |
| Akan | 1.00 [reference] |  | 1.00 [reference] |  |
| Ga/Dangme | 0.50 [0.36, 0.69] | <0.001 | 1.11 [0.74, 1.66] | 0.618 |
| Ewe | 0.64 [0.46, 0.88] | 0.007 | 1.33 [0.86, 2.08] | 0.203 |
| Guan | 0.97 [0.49, 1.93] | 0.931 | 2.10 [0.95, 4.64] | 0.067 |
| Mole Dagbani | 0.95 [0.69, 1.32] | 0.756 | 2.14 [1.35, 3.39] | 0.001 |
| Grusi | 0.55 [0.28, 1.06] | 0.076 | 1.07 [0.49, 2.33] | 0.864 |
| Gurma | 2.05 [1.65, 2.55] | <0.001 | 2.11 [1.31, 3.39] | 0.002 |
| Others | 1.06 [0.60, 1.88] | 0.837 | 2.47 [1.29, 4.74] | 0.006 |
| **Household size** |  |  |  | 0.026<sup>#</sup> |
| 1-3 members | 1.00 [reference] |  | 1.00 [reference] |  |
| 4-6 members | 0.72 [0.60, 0.86] | <0.001 | 1.08 [0.83, 1.40] | 0.565 |
| 7-9 members | 0.72 [0.57, 0.91] | 0.006 | 0.93 [0.66, 1.30] | 0.654 |
| 10+ members | 0.92 [0.69, 1.22] | 0.550 | 1.19 [0.80, 1.78] | 0.394 |
| Non-response | 0.48 [0.28, 0.80] | 0.005 | 0.44 [0.24, 0.79] | 0.006 |
| **Number of children** |  |  |  | 0.183<sup>#</sup> |
| No child | 1.00 [reference] |  | 1.00 [reference] |  |
| 1 child | 2.23 [1.76, 2.83] | <0.001 | 1.17 [0.83, 1.66] | 0.368 |
| 2-3 children | 2.15 [1.76, 2.61] | <0.001 | 1.44 [1.00, 2.06] | 0.048 |
| 4+ children | 2.12 [1.72, 2.62] | <0.001 | 1.20 [0.77, 1.86] | 0.414 |
| **Have any existing health condition** |  |  |  | 0.091<sup>#</sup> |
| No | 1.00 [reference] |  | 1.00 [reference] |  |
| Yes | 1.77 [1.48, 2.13] | <0.001 | 1.24 [0.97, 1.59] | 0.091 |
| **I believe CAMs are effective in preventing malaria.** |  |  |  | <0.001<sup>#</sup> |
| Agree | 1.00 [reference] |  | 1.00 [reference] |  |
| Neutral | 0.22 [0.17, 0.28] | <0.001 | 0.36 [0.25, 0.52] | <0.001 |
| Disagree | 0.35 [0.28, 0.43] | <0.001 | 0.72 [0.50, 1.04] | 0.081 |
| Non-response | 0.10 [0.03, 0.32] | <0.001 | 0.03 [0.00, 0.54] | 0.016 |
| **It is safe to use CAM in preventing malaria.** |  |  |  | 0.728<sup>#</sup> |
| Agree | 1.00 [reference] |  | 1.00 [reference] |  |
| Neutral | 0.34 [0.28, 0.42] | <0.001 | 1.00 [0.73, 1.37] | 0.996 |
| Disagree | 0.31 [0.25, 0.39] | <0.001 | 0.92 [0.61, 1.37] | 0.675 |
| Non-response | 0.31 [0.16, 0.63] | 0.001 | 2.33 [0.45, 11.99] | 0.310 |
| **Attitude towards CAM use** |  |  |  | <0.001<sup>#</sup> |
| Accepting | 1.00 [reference] |  | 1.00 [reference] |  |
| Ambivalent | 0.82 [0.67, 1.02] | 0.070 | 1.05 [0.77, 1.43] | 0.769 |
| Skeptical | 0.35 [0.26, 0.48] | <0.001 | 0.56 [0.37, 0.85] | 0.006 |
| Indifferent | 0.24 [0.19, 0.31] | <0.001 | 0.49 [0.35, 0.68] | <0.001 |
| **Perception about malaria:** |  |  |  |  |
| Consequence | 1.08 [1.05, 1.11] | <0.001 | 1.02 [0.97, 1.08] | 0.403 |
| Timeline | 1.02 [1.00, 1.05] | 0.069 |  |  |
| Personal control | 1.09 [1.06, 1.12] | <0.001 | 1.01 [0.97, 1.06] | 0.584 |
| Prevention/Treatment | 1.21 [1.18, 1.25] | <0.001 | 1.18 [1.12, 1.23] | <0.001 |
| Identity | 1.07 [1.04, 1.10] | <0.001 | 1.04 [0.98, 1.11] | 0.174 |
| Concerns | 0.98 [0.96, 1.01] | 0.236 |  |  |
| Understanding | 0.99 [0.96, 1.02] | 0.369 |  |  |
| Emotional response | 1.04 [1.01, 1.07] | 0.003 | 1.01 [0.96, 1.06] | 0.682 |

**Note:** COR=crude odds ratio; AOR=adjusted odds ratio; CI=confidence interval; #=Overall significance of variable
**Model Assessment:** Goodness of fit test p-value= 0.6445; Variance inflation Factor = 3.35; AUROCC [95% CI] = 0.8122 (CI: 0.7957, 0.8288)

Post estimation analysis showed that age group (p=0.003), highest education (p<0.001), employment status (p<0.001), ethnicity (p=0.008), household size (p=0.026), the believe that CAM is effective in preventing malaria (p<0.001), attitude of respondents towards CAM use (p<0.001) were overall significantly associated with the use of CAM in the last 12 months for malaria prevention.

## Discussion

This community-based cross-sectional study reports the use of CAM for malaria prevention in Ghana. Previous efforts and studies on CAM for malaria have mainly focused on treatment, with less attention to the preventative aspect. Although the literature often reports a high use of CAM for both the treatment and prevention of diseases in Ghana and across the continent, no study to date has reported on CAM use for malaria prevention. Our study aimed to estimate the prevalence, pattern, illness perception, and attitude of CAM for malaria prevention in the general population.

CAM use for malaria prevention in the last 12 months was 31.6%, suggesting that nearly one-third of the population engages in alternative approaches to reduce malaria risk. While no variations were noted between males and females in CAM use prevalence, significant differences in demographic characteristics were observed among the study participants. For example, significantly more males had higher education, were employed in the formal sector, and had fewer existing health conditions such as hypertension, peptic ulcer, and diabetes, compared with the female participants. Previously reported findings have, however, shown that CAM use in general was associated with being male [38], older (≥ 50 years), less educated, and unemployed or being employed in a blue-collar role [39]. These differences suggest that the determinants of CAM use for malaria prevention may be context-specific and influenced by both socioeconomic and health-related factors rather than sex alone.

Several malaria elimination strategies, including the widespread distribution and use of ITNs, indoor residual spraying, IPTp for pregnant women, and some herbal products for treating uncomplicated malaria, have significantly reduced malaria transmission. Despite these effective measures, many people continue to use herbal medicine and CAM to prevent diseases, including malaria, due to availability, accessibility, affordability, and cultural practices [25, 40, 41]. This is consistent with our findings, where biologically-based therapies, including botanical/herbal medicine, were the most utilized for malaria prevention. However, like with previous studies, CAM was used concurrently with orthodox medicines, and our study reported significant sex differences with this behaviour [22]. The concurrent use of CAM with orthodox medicines observed in this study may reflect a preference for a more holistic approach to health rather than a complete substitution of conventional care. However, this practice raises concerns about potential adverse effects and interactions between the two types of treatments [22]. The reported side effects, headaches, fatigue, and dizziness, highlight the need for closer monitoring and pharmacovigilance in the use of CAM products. [42, 43].

This study showed substantial regional variation in CAM use for malaria prevention, with the highest use in the Central region (55.6%) and lowest in the Greater Accra region (15.6%). CAM goods and services may be more affordable and accessible to people in relatively rural and socio-economically disadvantaged areas than in economically vibrant communities in the Greater Accra region.

Additionally, relative to participants with an accepting attitude, CAM use for malaria prevention was less among those who were skeptical and indifferent, which is consistent with previous studies [24, 37]. Generally, people are attracted to CAM products and services when they believe they are natural, safe, and have no unintended effects, thus influencing their accepting attitude towards CAM [44].

Perceptions of malaria significantly influence how people prevent or adapt to the illness [45]. These perceptions, formed through cognitive and emotional views about the disease, encompass anticipated physical, emotional, and social effects on the individual. Our study showed that, except for emotional response and treatment, a high median score was recorded for all other perception variables, including consequences (*i.e.,* belief about the effect of malaria), identity (*i.e.,* belief about malaria symptom experience), timeliness (*i.e.,* beliefs about the continuity of malaria), concerns (*i.e.,* beliefs about apprehension about malaria) and personal control (*i.e.,* beliefs about the uncontrollability of malaria through personal action) which showed a significant difference between males and females. The link between malaria risk perceptions and preventive behaviours has previously been reported [45]. Our result regarding the relationship between illness perceptions and CAM use for malaria prevention has the potential to help with malaria control and elimination strategies by identifying individual-level factors that could be potential targets for education and interventions.

The findings of the current study should be interpreted with caution considering several limitations. First, the cross-sectional design limits the ability to establish temporal or causal relationships between sociodemographic factors, perceptions, attitudes, and CAM use. Second, the main outcome was measured using self-reported data, which may be subject to recall bias and social desirability bias, potentially affecting the accuracy of the estimated prevalence of CAM use. Despite these limitations, the large sample size and multi-site design enhance the generalizability of the findings across diverse settings in Ghana.

Future research using more robust study designs, such as longitudinal or experimental studies, is urgently needed to assess the effectiveness and safety of CAM for malaria prevention. Such studies would provide stronger evidence to guide policy decisions and inform any potential integration of CAM with conventional malaria prevention strategies. Additionally, further research should explore the pharmacological properties of commonly used herbal products, potential drug-herb interactions, and the contextual factors influencing CAM use at the community level.

## Conclusion

Our findings revealed that about 32% of the population reported using CAM for malaria prevention in the last 12 months, highlighting its significant role in public health practices. The prevalence of CAM use did not differ between males and females. Preference for CAM, particularly botanical and herbal medicines, was associated with education, perceptions about malaria, employment, and attitudes about CAM. Thus, integrating CAM into malaria prevention strategies could enhance community acceptance and help with malaria elimination approaches. However, further research will be required to establish potential drug-herb interactions, validate the clinical efficacy of the herbal products, and isolate the lead compounds for an optimized malaria prevention therapy.

## Data Availability

All data produced in the present study are available upon reasonable request to the authors

## Abbreviations

aOR: Adjusted odds ratio
BMQ: Beliefs about Medicine Questionnaire
B-IPQ: Brief Illness Perception Questionnaire
CAM: Complementary and alternative medicine
CI: Confidence intervals
cORs: Crude odds ratios
FDA: Food and Drug Authority
IPTp: Intermittent preventive therapy in pregnant women
IPTi: Intermittent preventive therapy in infancy
IQR: Inter-quartile range
VIF: Variance inflation factor
WHO: World Health Organization

## Authors’ contributions

IAK, JPK, and KFMO contributed to the study conceptualization. IAK, AK, JPK, DA, YA, AFAM, MOA, and KFMO contributed to the study design and implementation. AK, DA, HKK, JAJ, NK, PTM, AM, AFAM, and MOA contributed to the study methodology and data collection. HAB and YA conducted data analysis, and IAK, YA, HAB, and KFMO contributed to data interpretation. IAK, JPK, YA, MOM and KFMO contributed to the draft. All authors contributed to the revision and critical evaluation of the manuscript for intellectual content, and they all agreed on the final version.

## Acknowledgments

We acknowledge the participants for their participation in this study.

## Availability of data and materials

The data will be shared on a reasonable request to the corresponding author.

## Ethics approval and consent to participate

The study was approved by the University of Ghana Medical Centre Institutional Review Board (UGMC-IRB), Legon, on August 30, 2023 (Protocol Number: UGMC-IRB/MSRC/033/2023). All study and data collection processes were performed per the Declaration of Helsinki in line with ensuring high ethical standards, providing information and obtaining consent from all participants, and data confidentiality. Participation was voluntary, and written informed consent was obtained from each participant before recruitment into the study.

## Consent for publication

Not applicable.

